# Clinical Meaningfulness of Individual Items of the Brief Ataxia Rating Scale and the Scale for the Assessment and Rating of Ataxia

**DOI:** 10.64898/2026.09.15.26362565

**Authors:** Defne Apaydin, Jeremy D. Schmahmann

**Affiliations:** Ataxia Center, Laboratory for Neuroanatomy and Cerebellar Neurobiology, Division of Behavioral Neurology, Department of Neurology, Massachusetts, General Hospital and Harvard Medical School, Boston, MA, USA

**Keywords:** Cerebellar disease, Spinocerebellar Ataxia, Cerebellar Motor Syndrome, Brief Ataxia Rating Scale, Scale for Assessment and Rating of Ataxia, Patient-Reported Outcome Measure of Ataxia, Natural History Study

## Abstract

**Introduction:** The Brief Ataxia Rating Scale (BARS) and Scale for the Assessment and Rating of Ataxia (SARA) are used to quantify cerebellar ataxia severity. We investigated how often clinicians’ ratings match patients’ reports of functional impairment, which scale items patients consider important, and whether incremental changes in item scores are meaningful to patients.

**Methods:** 30 patients with cerebellar ataxia completed BARS, SARA, FARS Functional Staging, and an Importance of Activities of Daily Living questionnaire. We studied BARS and SARA item scores for over- or underestimation in relation to patients’ perception of impairment. We explored patients’ rankings of item importance in relation to daily function, and their perception of the likely importance of 0.5- and 1-point changes in BARS gait and speech.

**Results:** Patients identified gait as most important on BARS and SARA, followed by oculomotor (BARS) and stance (SARA). Heel-to-shin was least important, with highest overestimation on both scales. Total overestimated points were similar between BARS (2.65 ± 2.80) and SARA (2.78 ± 2.78). 0.5-point changes in BARS gait and speech scores were rated very or extremely important by 60% and 46.6%, respectively, increasing to 83.3% and 70% for a 1-point change.

**Conclusion:** Gait is most meaningful to patients with cerebellar ataxia. BARS oculomotor and SARA stance rank second. BARS and SARA appendicular measures have limited patient-perceived clinical meaningfulness. 0.5-point increments in BARS gait and speech provide greater granularity of assessment and are considered meaningful by a substantial proportion of patients, supporting their relevance in practice and in clinical trials.

## 1. Introduction

The Brief Ataxia Rating Scale (BARS)^1^ and the Scale for the Assessment and Rating of Ataxia (SARA)^2^ have been extensively evaluated for reliability, validity, and sensitivity to change in patients with cerebellar disorders. We previously found a mosaic mapping between individual Clinical Outcome Assessment (COA) items and patient- and family-reported symptoms and impacts in daily life, demonstrating that BARS and SARA capture aspects of illness relevant to patients and families^3^. Here, we investigate how often a clinician’s rating over- or under-estimates patients’ reports of their functional impacts, which scale items patients consider most and least important, and whether incremental changes in these clinician-administered scales represent changes that patients perceive as meaningful.

## 2. Methods

### 2.1. Study Population

We recruited 30 patients: 13 from the National Ataxia Foundation (NAF) Clinical Research Consortium for the Study of Cerebellar Ataxia (CRC-SCA) at the Massachusetts General Hospital (MGH) Ataxia Center, 13 from the 2026 NAF Annual Ataxia Conference (AAC), 3 through participation in the Biohaven Troriluzole Expanded Access Program, and 1 from the MGH Ataxia Center. The 13 CRC-SCA patients completed their questionnaires by telephone 82.85 ± 74.72 days after their BARS and SARA assessments. The 13 AAC participants completed an Importance of Activities of Daily Living (IADL) questionnaire in person at the conference, and the remaining 4 patients completed the questionnaire in person.

### 2.2. Clinical Outcome Assessments (COAs)

BARS assesses ataxia severity across five tasks: gait; heel-to-shin (left and right); finger-to-nose (left and right); speech; and oculomotor, with a maximum score of 30. BARS version 2 uses a half-point grading system with each score explicitly detailed.

SARA assesses ataxia severity across eight tasks: gait, stance, sitting, speech, finger chase, finger-to-nose, fast alternating hand movements, and heel-to-shin, with appendicular items averaged across sides and a maximum score of 40.

*Friedreich’s Ataxia Rating Scale (FARS) Functional Staging*^4^ (FARS-Func) is a single-item subscale, 0 to 6, assessing overall functional status. We used FARS-Func to classify patients’ ataxia severity into mild: normal or no disability (Stages < 2.0); moderate: minimal disability, use of a cane or holding onto walls or furniture (Stages ≥ 2.0 and < 4.0); and severe: requires a walker or is confined to a wheelchair (Stages ≥ 4.0).

The IADL questionnaire adapted from Maas et al.^5^ comprises 10 yes/no questions assessing functional difficulty, mapped to corresponding BARS and SARA items: walking (gait); balance while standing (stance); balance while sitting unsupported (sitting); putting on socks and/or shoes and placing and keeping the feet on the pedals while driving (heel-to-shin); fine motor control, such as writing, buttoning clothes, and putting on jewelry (finger-to-nose, finger chase, and fast alternating hand movements); speech (speech); and vision, reading ability, and visual difficulties while driving (oculomotor). Patients identified their most troublesome disease-related symptoms, the symptom they would most like to see improved, and the most and least important items on BARS and SARA. Because BARS uses 0.5-point increments, we evaluated the perceived meaningfulness of hypothetical 0.5- and 1-point improvements or declines from each patient’s current score for gait and speech, but not finger-to-nose and heel-to-shin, because their technical descriptions are less interpretable for real-world function. The definitions corresponding to the patient’s current score were read aloud, followed by those corresponding to scores reflecting 0.5-point and 1-point changes. Patients rated the importance of each hypothetical change on a 5-point Likert scale (0 = not important at all, 1 = slightly important, 2 = moderately important, 3 = very important, 4 = extremely important).

### 2.3. Data Analysis

Data analysis and visualization were conducted in R. Concordance between scale scores, and patient-reported impairment was classified as overestimation, underestimation, or match. Overestimation was defined as BARS or SARA item score > 0 in the absence of corresponding IADL complaint; underestimation as item score 0 despite corresponding IADL complaint; and a match was concordance between clinician-rated item score and patient’s corresponding IADL response. Number of overestimated points for BARS and SARA were calculated as the sum of item scores classified as overestimation for each patient. Mean overestimated points were compared using a paired Wilcoxon signed-rank test. Spearman’s correlation coefficient was used to assess associations between BARS gait and speech scores, FARS-Func, and 0.5- and 1-point importance ratings, with p-values adjusted for multiple comparisons using a Holm correction.

## 3. Results

30 participants (17 females; age 54.17 ± 14.94 years) completed the study. 10 were mild, 14 moderate, and 6 severe based on FARS-Func. The cohort comprised 12 spinocerebellar ataxia type 3 (SCA3), 7 SCA27B, 3 SCA2, 2 SCA1, and 2 SCA6 patients, and single patients each contributing SCA7, SCA8, cerebellar ataxia with neuropathy and vestibular areflexia syndrome (CANVAS), and an autoimmune-mediated cerebellar syndrome.

### 3.1. Meaningfulness to patients of individual items

On BARS, 21 patients (70%) selected gait as the most important item, followed by oculomotor function (16.7%), speech (10%), and finger-to-nose (3.3%) (Figure 1A). Five patients (16.7%) reported the oculomotor item on BARS to be the most important, of whom three had SCA27B (Figure 1A). On SARA, 23 patients (76.7%) selected gait as the most important, followed by stance (13.3%), speech (6.7%), and finger chase (3.3%) (Figure 1B). Heel-to-shin was selected as least important on both scales, 43.3% of patients on BARS and 40% on SARA; finger-to-nose least important in 30% of patients on BARS and 13.3% on SARA; and finger chase least important in 16.7% on SARA (Figures 1C and 1D). Gait and stance were the symptoms that patients most wanted to improve (Figures 1E and 1F). Other troubling symptoms reported by patients are in Figures 1E and 1F.

**Figure 1.**
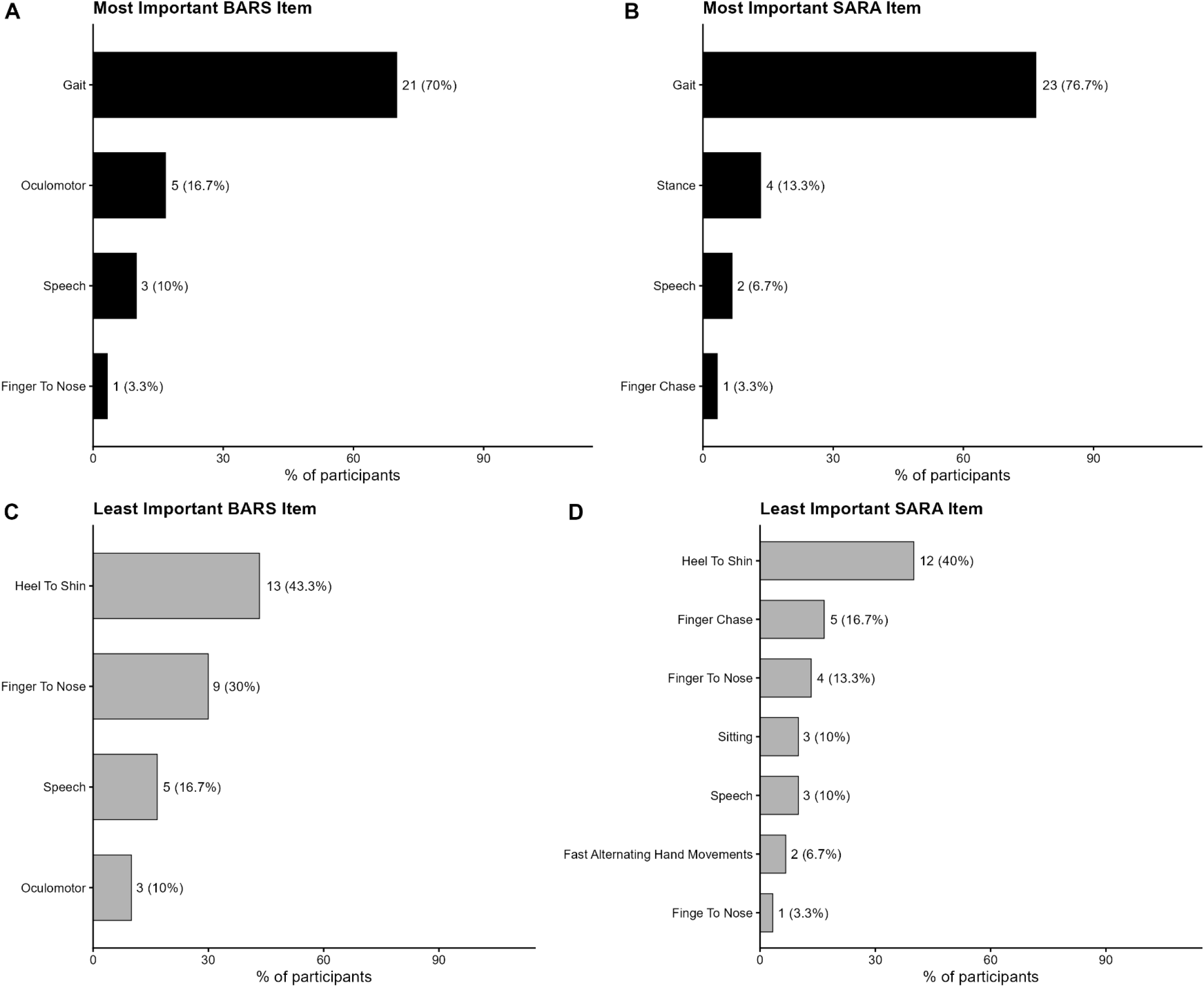
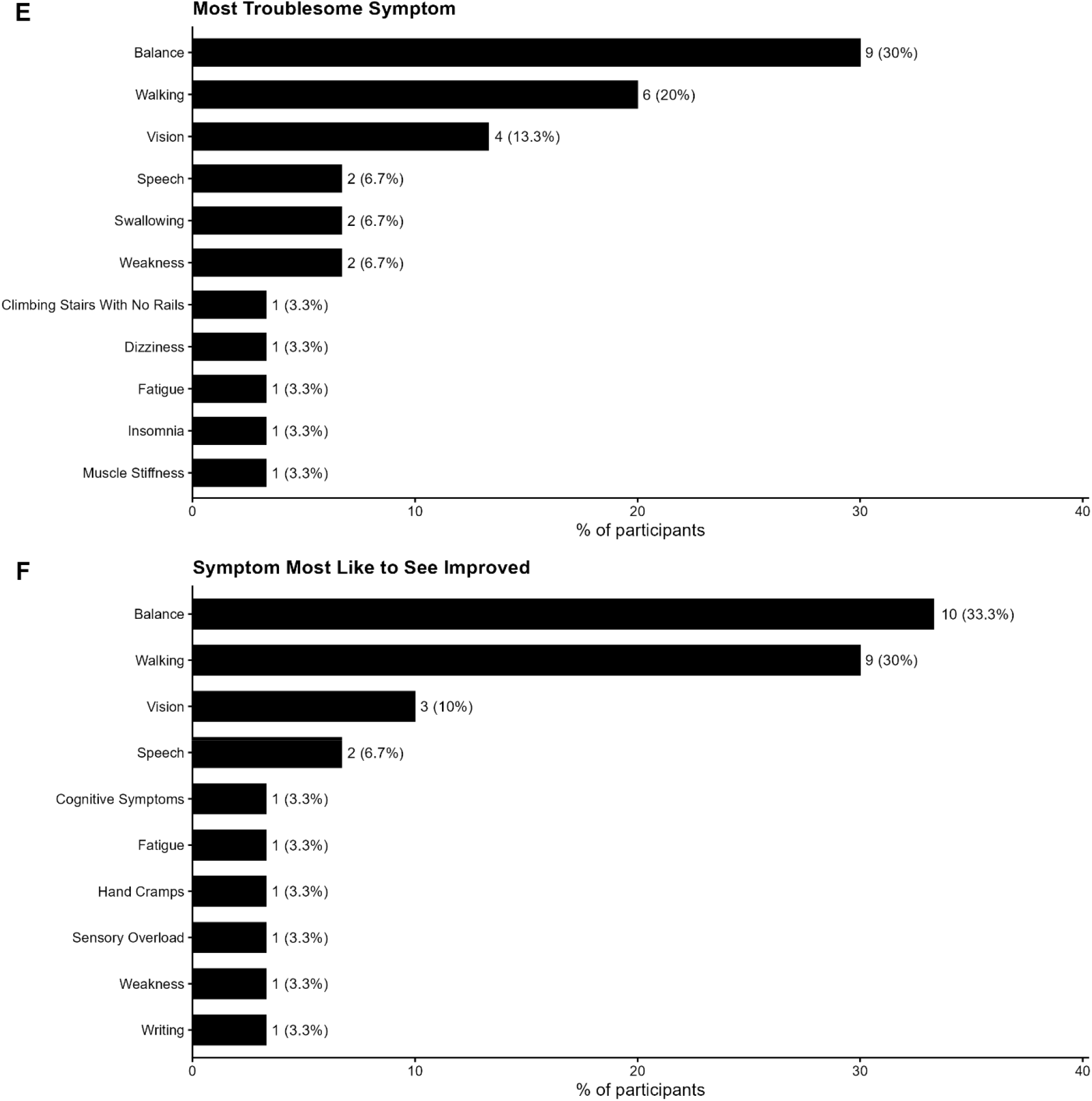
**A-D**. Patient-reported item importance rankings for the BARS and SARA scales. Horizontal bars show the percentage of the cohort (N = 30) identifying each item as the most important item or least important. Panels A and B show the mo in the BARS (top row) and SARA (bottom row) rating scales. **Figure 1E-F**. Patient-reported most troublesome symptom and symptom most like to see improved. Horizontal bars show the percentage of the cohort (N = 30) reporting their most troublesome symptom (top), and the symptom they would most like to see improved (bottom).

### 3.2. Meaningfulness to patients of potential change in BARS gait and speech scores

60% of patients rated a 0.5-point change in BARS gait as very or extremely important (43.3% and 16.7%, respectively), increasing to 83.3% for a 1.0-point change (40.0% and 43.3%, respectively).

A 0.5-point change in BARS speech was rated as very or extremely important by 46.6% of patients (33.3% and 13.3%, respectively), increasing to 70.0% for a 1.0-point change (43.3% and 26.7%, respectively; Figures 2A-D). Higher BARS speech scores were associated with greater importance for a 0.5-point change (ρ = 0.48, Holm-adjusted p = 0.03) (Supplementary Figure S1G).

**Figure 2.**
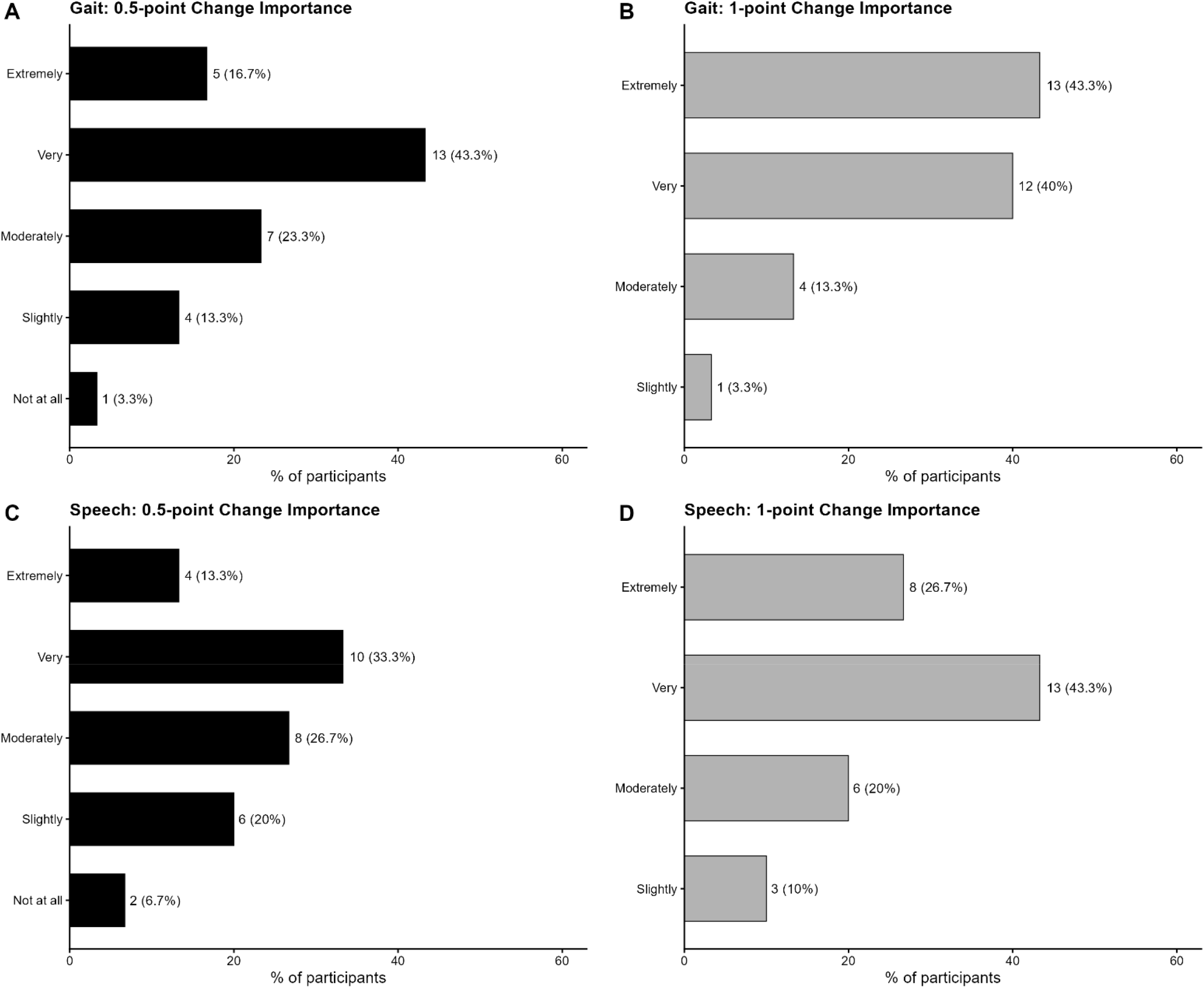
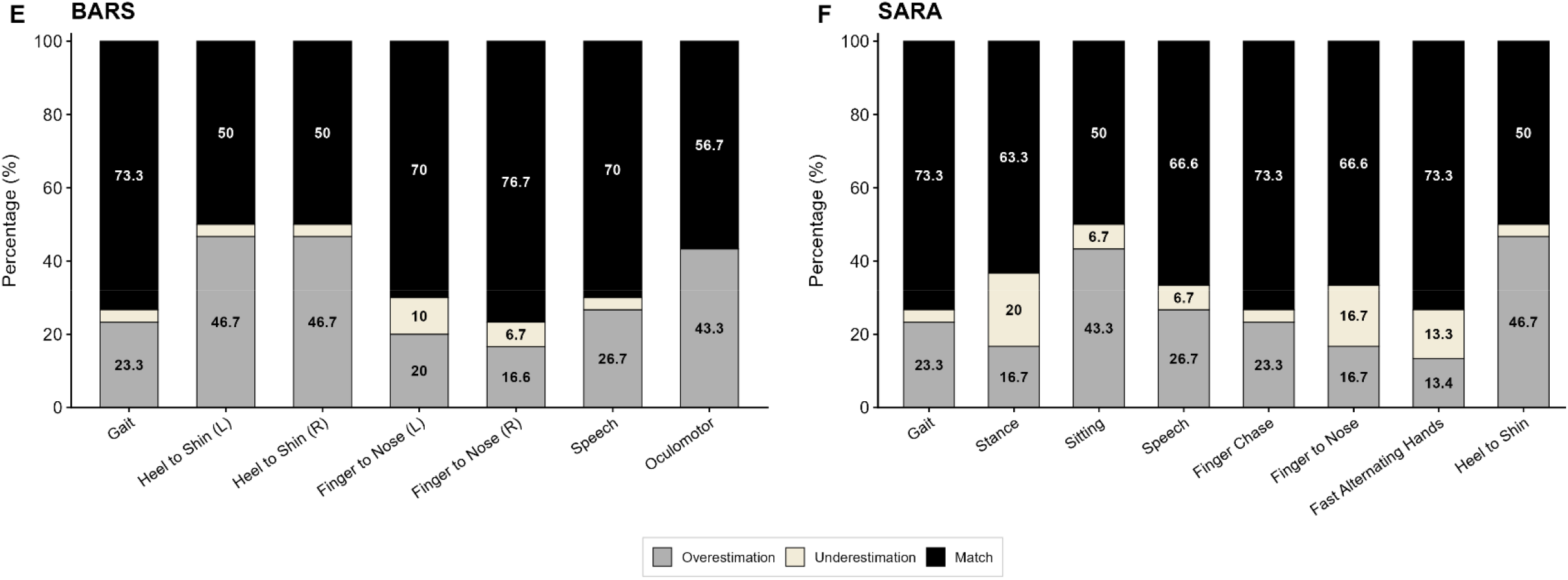
**A-B**. Distribution of patient-reported importance ratings for 0.5-point and 1-point changes in BARS gait and speech scores. Horizontal bars show the percentage of the cohort (N = 30) selecting each importance rating (Not at all, Slightly, Moderately, Very, and Extremely Important) when asked how much a 0.5-point or 1-point change in their BARS gait or speech score would matter to them. **Figure 2C-F**. Discrepancy between BARS and SARA item scores and patient-reported activities of daily living complaints. For each BARS and SARA item, stacked bars show the percentage of the cohort (N = 30) classified as overestimation (item score > 0 with no corresponding complaint on the IADL), underestimation (item score = 0 despite a corresponding complaint on the IADL), or match (item score and IADL complaint are in agreement).

### 3.3. Item rating and symptom concordance

On BARS, gait, finger-to-nose, and speech showed fair concordance. Heel-to-shin and oculomotor items were the most overestimated (46.7% and 43.3%; Figure 2E). On SARA, heel-to-shin was the most overestimated (46.7%), followed by sitting (43.3%; Figure 2F). Mean total number overestimated points was 2.65 ± 2.80 (BARS) and 2.78 ± 2.78 (SARA), with no significant difference between them (Wilcoxon signed-rank test, *V* = 199.5, p =0.6). See Supplementary Table 1 for individual BARS and SARA scores.

## 4. Discussion

In this study we explored alignment between clinician-rated impairment on BARS and SARA and patient-reported difficulty on ADLs in patients with cerebellar ataxia, the meaningfulness of the individual items on the COAs for patients, and the perceived meaningfulness of incremental BARS gait and speech score changes (0.5 and 1.0 points).

### 4.1. Meaningfulness to patients of individual items

The key finding was that patients with ataxia considered gait to be by far the single most important item in both COAs. Oculomotor (BARS) and stance (SARA) were ranked similarly. Speech was ranked next in importance on both scales, while upper extremity measures were ranked low, and heel-to-shin ranked lowest.

### 4.2. Meaningfulness to patients of potential change in BARS gait and speech scores

There was a high proportion of patients who rated a 0.5-point change in BARS gait and speech as very or extremely important, and for a 1-point change this was even more prominent. 0.5-point speech changes were more meaningful in patients with higher BARS speech scores, suggesting that speech may assume greater importance as dysarthria becomes more severe.

### 4.3. Item rating and symptom concordance

The most overestimated item on both BARS and SARA (COA score > patient report) was the heel-to-shin test, suggesting that poor performance on heel-to-shin may not translate into meaningful functional limitations. Maas et al.^5^ did not include a heel-to-shin-related ADL item in their questionnaire, noting that no ADL involves isolated lower-limb coordination beyond walking and standing. In an attempt to address this and explore the potential daily-life relevance of the heel-to-shin test, we included two additional questions: “*Do you have any difficulties putting on socks/shoes?” and “Do you have any difficulty with placing and keeping your feet on the pedals while driving?*”. We found that, whereas the test may provide useful information for clinicians about a patient’s disease severity it did not necessarily translate into meaningful impairment in these specific activities.

The second most overestimated BARS item was oculomotor function. We note that patients with SCA27B reported greater functional relevance of oculomotor abnormalities, consistent with the vestibular basis of their gait disorder. Of the 13 out of 30 patients (43.3%) whose oculomotor impairment was overestimated, only two had SCA27B, one of whom was receiving dalfampridine which may have mitigated the functional impact of the impairment, and medication data were unavailable for the other. Oculomotor abnormalities, including saccadic pursuit, hypermetric saccades, downbeat nystagmus in primary gaze, gaze-evoked downbeat and/or direction-beating nystagmus, symptomatic diplopia, and visual flow phenomena, are core features of cerebellar ataxia^6^ and are particularly prominent in SCA27B^7^, which accounts for ~16% of inherited SCAs^8^. Hirschfeld et al.^9^ reported saccadic pursuit in 80.7% of patients with cerebellar ataxia, and nystagmus in 71.2%, and we previously showed that oculomotor abnormalities are ubiquitous in the SCAs and identifiable in nearly all patients at their initial visit^10^. In this context it is notable that 16.7% of our cohort considered BARS oculomotor the most important item on the scale.

Gait was overestimated in 7 patients in whom the highest score was BARS 1.5 (Cadence, speed, stance is slightly irregular, may take an extra step to turn but not clearly abnormal) and SARA 2 (Clearly abnormal, tandem walking > 10 steps not possible). These mildly affected individuals may not perceive subtle gait abnormalities as meaningful functional limitations, despite the examination finding of difficulty with tandem walking.

The mean total number of overestimated points was similar for BARS (2.65 ± 2.80) and SARA (2.78 ± 2.78), as also seen in Maas et al.^5^ The lack of statistically significant difference between BARS and SARA suggests that this finding is not unique to either scale but may reflect a limitation shared by both COAs. BARS and SARA are designed to quantify observable impairments, but we show that not all items correspond to patients’ perceptions of their functional limitations.

In sum, in this cohort we report that (i) gait was the domain considered most important by patients, followed by oculomotor and stance; (ii) a 0.5-point change in BARS gait and speech was considered meaningful by a substantial proportion of patients, thus supporting the clinical relevance of measuring changes of this small magnitude, and (iii) appendicular measures were overestimated by the COAs compared with patient-reported functional impairment.

## 5. Limitations

There was a variable delay between BARS/SARA administration and completion of the IADL questionnaire by CRC-SCA participants (82.85 ± 74.72 days). The expected progression in ataxia severity over this relatively short interval is small^11^, making it unlikely that this delay substantially affected questionnaire responses. Our sample size of 30 was modest but appropriate for this study which was designed to characterize patient perceptions of individual scale items and incremental score changes and to explore their relationship to patient-reported functional difficulty.

## 6. Conclusion

What clinicians measure is not always what patients experience as functionally meaningful. This has implications for which components of ataxia rating scales, and what magnitude of change, should be emphasized in clinical trials. We found that 0.5-point increments in BARS gait and speech enhance granularity of assessment, and these small increments are considered clinically meaningful by a substantial proportion of patients. The BARS oculomotor and SARA stance items rank second to gait in importance to patients. Appendicular measures are overestimated by both BARS and SARA and have limited patient-perceived clinical meaningfulness. These findings have implications for the construct and interpretation of clinical outcome assessments used in ataxia clinical trials.

## Data Availability

All data produced in the present work are contained in the manuscript

## Ethical Compliance Statement

This study was approved by the Mass General Brigham Institutional Review Board (IRB) on March 18th, 2026 (IRB number 2026P000653). Informed consent was not required for this study. As part of the recruitment process, patients were provided with a fact sheet describing the study. Patients provided implied consent by choosing to participate or decline the invitation. We confirm that we have read the Journal’s position on issues involved in ethical publication and affirm that this work is consistent with those guidelines.

## CRediT authorship contribution statement

**Defne Apaydin:** Methodology, Data curation, Formal analysis, Investigation, Visualization, Writing – original draft, Writing – review & editing. **Jeremy Schmahmann:** Conceptualization, Methodology, Formal Analysis, Investigation, Resources, Funding acquisition, Project administration, Supervision, Validation, Writing – original draft, review & editing.

