## Supplementary Figures for "Clinical Meaningfulness of Individual Items of the Brief Ataxia Rating Scale and the Scale for the Assessment and Rating of Ataxia"

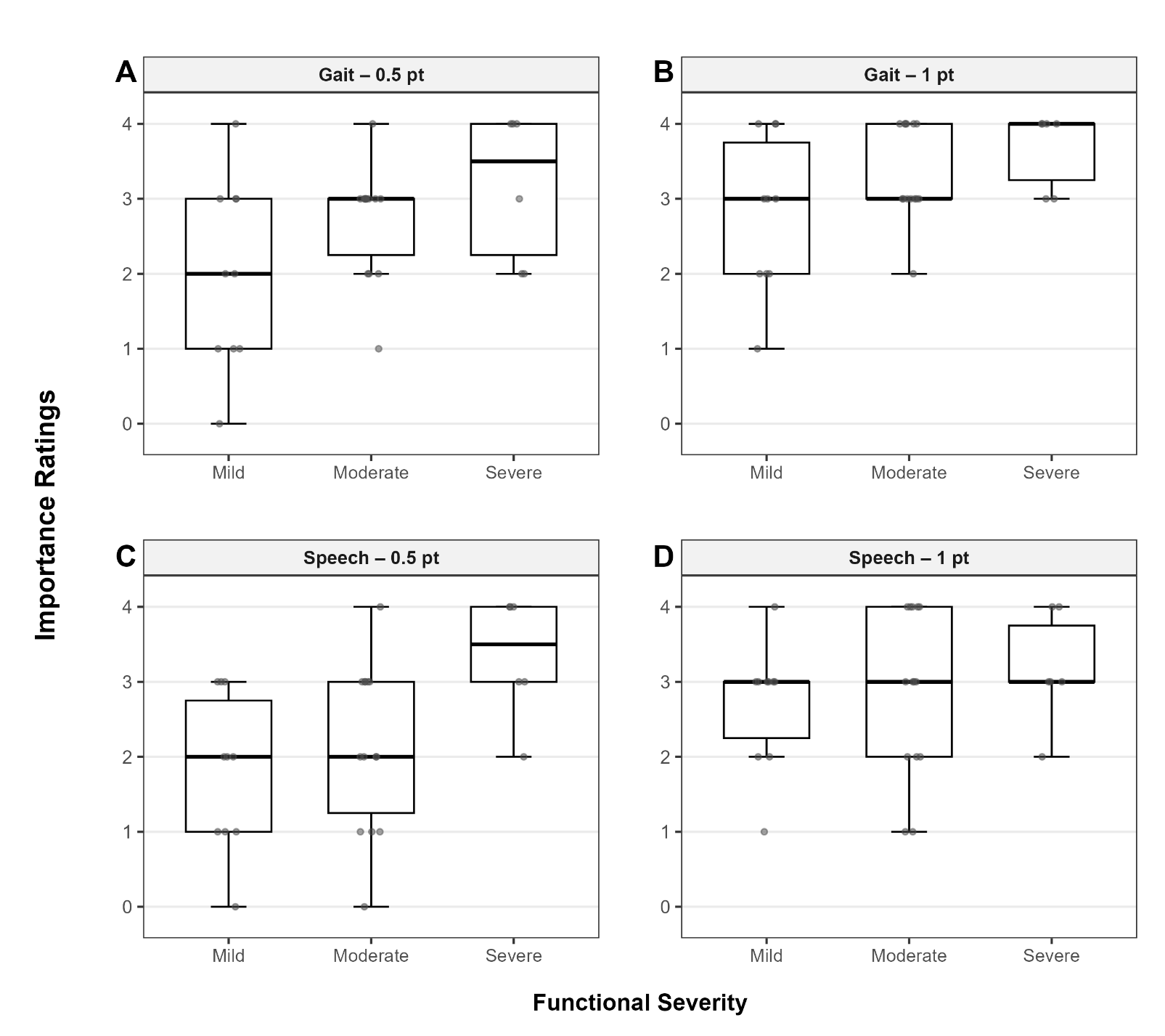

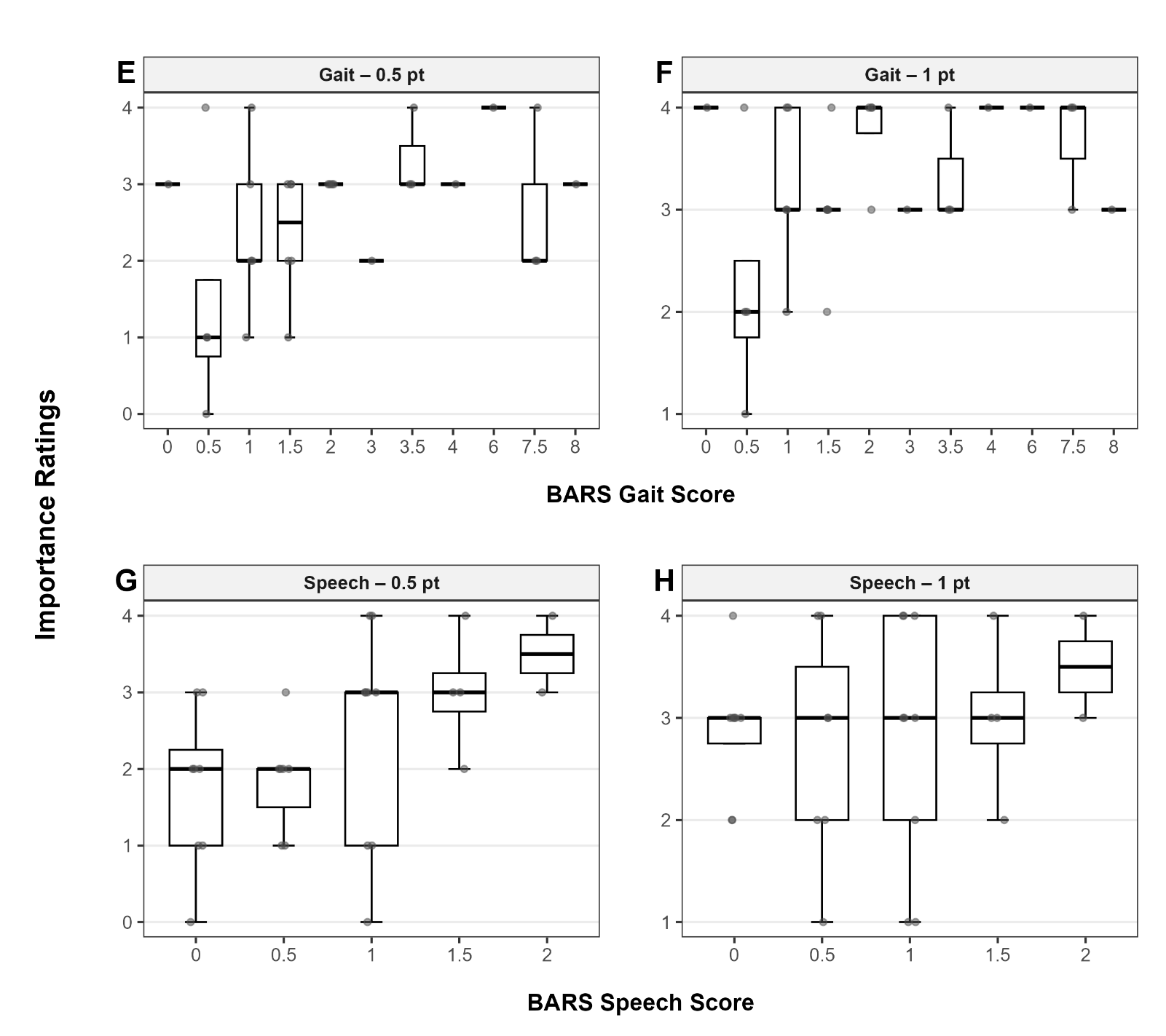

**Figures S1A-D.** Patient-reported importance of BARS gait and speech score changes, grouped by Functional Severity measured by the Friedreich Ataxia Rating Scale Functional Staging. Box-and-whisker plots show the distribution of importance ratings (0 = Not at all to 4 = Extremely) for a 0.5-point or 1-point change in BARS gait or speech scores, grouped by FARS Functional Stage severity band (Mild, Moderate, Severe; corresponding raw FARS Functional Stage scores shown beneath each category). Points represent individual patients; the horizontal line within each box indicates the group median; boxes represent the interquartile range. **Figures S1E-H.** Patient-reported importance of BARS Gait and Speech score changes, grouped by current BARS domain score. Box-and-whisker plots show the distribution of importance ratings (0 = Not at all to 4 = Extremely) for a 0.5-point or 1-point change in BARS gait and speech scores. Points represent individual patients; the horizontal line within each box indicates the group median; boxes represent the interquartile range. Patients with more impairment in speech tended to report a 0.5-point change in speech severity as more meaningful (ρ = 0.48, Holm-adjusted p = 0.03).
