## Supplementary Table for "Clinical Meaningfulness of Individual Items of the Brief Ataxia Rating Scale and the Scale for the Assessment and Rating of Ataxia"

**Table S1A.** BARS item and total scores by patient. To protect patient privacy, age is reported as non-overlapping quartile ranges spanning the cohort's age range, each 13 years wide: 28–40, 41–53, 54–66, and 67–79 years.

| **ID** | **Age**  **(y)** | **Sex** | **Diagnosis** | **Gait** | **Heel to**  **Shin (L)** | **Heel to**  **Shin (R)** | **Finger to**  **Nose (L)** | **Finger to**  **Nose (R)** | **Speech** | **Oculomotor** | **Total**  **BARS Score** |
| --- | --- | --- | --- | --- | --- | --- | --- | --- | --- | --- | --- |
| 1 | 28–40 | F | SCA2 | 1.5 | 2.5 | 1.5 | 2 | 1.5 | 1 | 0.5 | 10.5 |
| 2 | 67–79 | M | SCA3 | 7.5 | 2 | 2.5 | 1 | 1.5 | 2 | 1.5 | 18 |
| 3 | 28–40 | F | SCA3 | 0.5 | 1 | 0.5 | 0 | 0 | 0 | 1 | 3 |
| 4 | 28–40 | M | SCA1 | 1 | 2.5 | 1.5 | 1 | 0.5 | 1 | 1.5 | 9 |
| 5 | 67–79 | F | SCA6 | 7.5 | 2.5 | 1.5 | 1.5 | 1.5 | 1 | 1 | 16.5 |
| 6 | 67–79 | F | SCA27B | 0.5 | 1 | 0.5 | 0.5 | 0.5 | 0 | 1.5 | 4.5 |
| 7 | 67–79 | F | SCA27B | 1 | 2 | 1 | 1.5 | 1 | 0.5 | 1.5 | 8.5 |
| 8 | 28–40 | F | SCA3 | 1.5 | 1.5 | 1 | 0.5 | 0.5 | 1 | 1.5 | 7.5 |
| 9 | 41–53 | F | SCA3 | 0.5 | 1.5 | 1 | 0 | 0 | 0 | 1 | 4 |
| 10 | 28–40 | M | SCA3 | 2 | 2 | 2 | 1 | 1 | 0.5 | 1 | 9.5 |
| 11 | 41–53 | M | SCA8 | 7.5 | 3 | 2.5 | 2 | 2 | 1.5 | 1.5 | 20 |
| 12 | 54–66 | F | SCA1 | 3 | 2 | 2.5 | 1 | 1 | 1 | 2 | 12.5 |
| 13 | 67–79 | F | SCA6 | 1.5 | 0 | 0 | 0.5 | 0 | 0.5 | 1 | 3.5 |
| 14 | 28–40 | F | Autoimmune mediated cerebellar syndrome | 1.5 | 3.5 | 3 | 1 | 0.5 | 1 | 0.5 | 11 |
| 15 | 54–66 | F | SCA7 | 4 | 3 | 2.5 | 1.5 | 1 | 1.5 | 1 | 14.5 |
| 16 | 28–40 | F | SCA2 | 8 | 3 | 3 | 2 | 2 | 1.5 | 1 | 20.5 |
| 17 | 41–53 | M | SCA2 | 2 | 1 | 1 | 0.5 | 0.5 | 0.5 | 0.5 | 6 |
| 18 | 54–66 | F | SCA3 | 3.5 | 2.5 | 1.5 | 1 | 1 | 1 | 1 | 11.5 |
| 19 | 54–66 | M | SCA3 | 1.5 | 2.5 | 2.5 | 1 | 1 | 0.5 | 1.5 | 10.5 |
| 20 | 67–79 | F | SCA27B | 1 | 1 | 0.5 | 0 | 0 | 0 | 1 | 3.5 |
| 21 | 41–53 | M | SCA3 | 1.5 | 0 | 0.5 | 0 | 0 | 0 | 1.5 | 3.5 |
| 22 | 54–66 | M | SCA27B | 6 | 0.5 | 0.5 | 1 | 1 | 2 | 1.5 | 12.5 |
| 23 | 67–79 | M | SCA27B | 0 | 0 | 0 | 0.5 | 0.5 | 0.5 | 1 | 2.5 |
| 24 | 41–53 | F | SCA3 | 3.5 | 1.5 | 1.5 | 1 | 1 | 1 | 1 | 10.5 |
| 25 | 54–66 | M | CANVAS | 3.5 | 2.5 | 2.5 | 1.5 | 1 | 1.5 | 2 | 14.5 |
| 26 | 41–53 | M | SCA3 | 2 | 1 | 2 | 0.5 | 0.5 | 1 | 1.5 | 8.5 |
| 27 | 41–53 | M | SCA3 | 1 | 1 | 1.5 | 0 | 0.5 | 0.5 | 1 | 5.5 |
| 28 | 28–40 | F | SCA3 | 0.5 | 0 | 0 | 0 | 0 | 0 | 0.5 | 1 |
| 29 | 67–79 | F | SCA27B | 2 | 0 | 0 | 0.5 | 0.5 | 0 | 1.5 | 4.5 |
| 30 | 67–79 | M | SCA27B | 1 | 0.5 | 0 | 0 | 0 | 0 | 1.5 | 3 |

**Table S1B.** SARA item and total scores by patient. To protect patient privacy, age is reported as non-overlapping quartile ranges spanning the cohort's age range, each 13 years wide: 28–40, 41–53, 54–66, and 67–79 years.

| **ID** | **Age**  **(y)** | **Sex** | **Diagnosis** | **Gait** | **Stance** | **Sitting** | **Speech** | **Finger**  **Chase** | **Finger to**  **Nose** | **Alternating**  **Hand**  **Movements** | **Heel to**  **Shin** | **Total**  **SARA Score** |
| --- | --- | --- | --- | --- | --- | --- | --- | --- | --- | --- | --- | --- |
| 1 | 28–40 | F | SCA2 | 2 | 0 | 1 | 2 | 1 | 1.5 | 0.5 | 1.5 | 9.5 |
| 2 | 67–79 | M | SCA3 | 7 | 3 | 2 | 3 | 1 | 2 | 2.5 | 2.5 | 23 |
| 3 | 28–40 | F | SCA3 | 1 | 0 | 0 | 0 | 0.5 | 0 | 0.5 | 1 | 3 |
| 4 | 28–40 | M | SCA1 | 2 | 2 | 1 | 2 | 2 | 1.5 | 2 | 1.5 | 14 |
| 5 | 67–79 | F | SCA6 | 6 | 1 | 0 | 2 | 1 | 2 | 2.5 | 2.5 | 17 |
| 6 | 67–79 | F | SCA27B | 1 | 0 | 0 | 0 | 1.5 | 1 | 0 | 1 | 4.5 |
| 7 | 67–79 | F | SCA27B | 2 | 1 | 1 | 1 | 1 | 2 | 1 | 1.5 | 10.5 |
| 8 | 28–40 | F | SCA3 | 2 | 2 | 1 | 2 | 1.5 | 1 | 1.5 | 1.5 | 12.5 |
| 9 | 41–53 | F | SCA3 | 1 | 2 | 1 | 0 | 0.5 | 0 | 1 | 1 | 6.5 |
| 10 | 28–40 | M | SCA3 | 2 | 2 | 0 | 1 | 0 | 1 | 1 | 2.5 | 9.5 |
| 11 | 41–53 | M | SCA8 | 6 | 5 | 2 | 2 | 2.5 | 2.5 | 3 | 2.5 | 25.5 |
| 12 | 54–66 | F | SCA1 | 3 | 2 | 1 | 2 | 1.5 | 2 | 2.5 | 2 | 16 |
| 13 | 67–79 | F | SCA6 | 2 | 0 | 0 | 1 | 1 | 0 | 0 | 0 | 4 |
| 14 | 28–40 | F | Autoimmune mediated cerebellar syndrome | 2 | 1 | 0 | 2 | 1.5 | 0.5 | 1 | 3 | 11 |
| 15 | 54–66 | F | SCA7 | 4 | 2 | 2 | 2 | 1 | 1.5 | 3 | 3 | 18.5 |
| 16 | 28–40 | F | SCA2 | 8 | 6 | 2 | 2 | 1 | 2 | 3 | 3 | 27 |
| 17 | 41–53 | M | SCA2 | 2 | 1 | 0 | 1 | 1 | 1 | 1.5 | 1 | 8.5 |
| 18 | 54–66 | F | SCA3 | 4 | 2 | 1 | 2 | 1.5 | 1 | 0 | 1 | 12.5 |
| 19 | 54–66 | M | SCA3 | 2 | 2 | 1 | 1 | 1 | 1 | 1 | 2.5 | 11.5 |
| 20 | 67–79 | F | SCA27B | 2 | 0 | 0 | 0 | 0 | 0 | 0 | 0.5 | 2.5 |
| 21 | 41–53 | M | SCA3 | 2 | 0 | 0 | 0 | 1 | 0 | 0 | 0.5 | 3.5 |
| 22 | 54–66 | M | SCA27B | 6 | 3 | 1 | 3 | 1 | 1 | 2 | 1 | 18 |
| 23 | 67–79 | M | SCA27B | 0 | 0 | 0 | 0 | 0.5 | 0 | 0 | 0 | 0.5 |
| 24 | 41–53 | F | SCA3 | 4 | 3 | 2 | 2 | 1 | 1 | 1 | 3 | 17 |
| 25 | 54–66 | M | CANVAS | 4 | 3 | 1 | 2 | 2 | 1 | 1 | 2 | 16 |
| 26 | 41–53 | M | SCA3 | 2 | 2 | 0 | 2 | 2 | 0.5 | 0.5 | 1.5 | 10.5 |
| 27 | 41–53 | M | SCA3 | 2 | 0 | 0 | 1 | 0.5 | 0 | 0 | 2 | 5.5 |
| 28 | 28–40 | F | SCA3 | 1 | 0 | 0 | 0 | 0 | 0 | 0 | 0 | 1 |
| 29 | 67–79 | F | SCA27B | 2 | 2 | 1 | 0 | 0 | 0 | 0 | 0 | 5 |
| 30 | 67–79 | M | SCA27B | 1 | 0 | 0 | 0 | 1 | 0 | 0 | 0 | 2 |
